# Neural Mechanisms of Acceptance and Commitment Therapy for Chronic Pain: A Network-Based fMRI Approach

**DOI:** 10.1101/2020.08.10.20144063

**Authors:** Semra A. Aytur, Kimberly L. Ray, Sarah K. Meier, Jenna Campbell, Barry Gendron, Noah Waller, Donald A. Robin

## Abstract

Over 100 million Americans suffer from chronic pain (CP), which causes more disability than any other medical condition in the U.S. at a cost of $560-$635 billion per year (IOM, 2011). Opioid analgesics are frequently used to treat CP. However, long term use of opioids can cause brain changes such as opioid-induced hyperalgesia that, over time, increase pain sensation. Also, opioids fail to treat complex psychological factors that worsen pain-related disability, including beliefs about and emotional responses to pain. Cognitive behavioral therapy (CBT) can be efficacious for CP. However, CBT generally does not focus on important factors needed for long-term functional improvement, including attainment of personal goals and the psychological flexibility to choose responses to pain.

Acceptance and Commitment Therapy (ACT) has been recognized as an effective, non-pharmacologic treatment for a variety of CP conditions. However, little is known about the neurologic mechanisms underlying ACT. We conducted an ACT intervention in women (n=9) with chronic musculoskeletal pain. Functional magnetic resonance imaging (fMRI) data were collected pre- and post-ACT, and changes in functional connectivity (FC) were measured using Network-Based Statistics (NBS). Behavioral outcomes were measured using validated assessments such as the Acceptance & Action Questionnaire (AAQ-II), the Chronic Pain Acceptance Questionnaire (CPAQ), the Center for Epidemiologic Studies Depression Scale (CES-D), and the NIH Toolbox Neuro-QoL™ (Quality of Life in Neurological Disorders) scales.

Results suggest that, following the four-week ACT intervention, participants exhibited reductions in brain activation within and between key networks including self-reflection (default mode, DMN), emotion (salience, SN), and cognitive control (frontal parietal, FPN). These changes in connectivity strength were correlated with changes in behavioral outcomes including decreased depression and pain interference, and increased participation in social roles. This study is one of the first to demonstrate that improved function across the DMN, SN, and FPN may drive the positive outcomes associated with ACT. This study contributes to the emerging evidence supporting the use of neurophysiological indices to characterize treatment effects of alternative and complementary mind-body therapies.

**Perspective:** This article identifies neural mechanisms that may mediate behavioral changes associated with Acceptance and Commitment Therapy (ACT) in persons with chronic musculoskeletal pain. This information could potentially help clinicians to determine which mind-body therapies may benefit specific patients as part of an integrative pain management approach.

## Introduction

Over 100 million Americans suffer from chronic pain (CP), which causes more disability than any other medical condition in the U.S. at a cost of $560-$635 billion per year (IOM, 2011). Opioid analgesics are frequently used to treat CP. However, long term use of opioids can cause brain changes such as opioid-induced hyperalgesia that, over time, increase pain sensation. Also, opioids fail to treat complex psychological factors that worsen pain-related disability, including beliefs and emotional responses to pain. Cognitive behavioral therapy (CBT) can be efficacious for CP (Lim et al., 2018). However, CBT does not focus on important factors needed for long-term functional improvement, including attainment of personal goals and the psychological flexibility to choose responses to pain (Wetherell et al., 2011).

Acceptance and Commitment Therapy (ACT) is a mindfulness-based therapy that focuses on enabling individuals to accept what is out of their control, and to commit to valued actions that enrich their lives (Vowles & McCracken, 2008). ACT was developed in 1986 by Stephen C. Hayes who began to examine how language and thought influence internal experiences (Harris, 2006). By emphasizing acceptance instead of avoidance, ACT differs from many other forms of cognitive behavioral therapy. Although not originally designed for CP, ACT has been shown to be efficacious in terms of clinical outcomes, adherence to treatment, and retention, earning the status of a “well-established” treatment for CP from the American Psychological Association. ACT aims to increase *psychological flexibility*, and has been associated with improved health outcomes in many randomized controlled clinical trials (Feliu-Soler et al., 2018), including three systematic reviews specific to CP (Hann et al., 2014; Veehof et al., 2016; Hughes et al., 2017). Psychological flexibility is defined as an individual’s ability to “recognize and adapt to various situational demands; shift mindsets or behavioral repertoires when these strategies compromise personal or social functioning; maintain balance among important life domains; and be aware, open, and committed to behaviors that are congruent with deeply held values” (Kashdan & Rottenberg, 2010; p865). ACT is a “third wave” behavioral treatment that has been shown to be efficacious for treating CP, as well as co-morbid conditions and factors (e.g., goal selection) related to long-term functional improvement (Vowles & McCracken, 2008; Yu & McCracken, 2017). Additionally, patients who participate in ACT report greater long-term satisfaction compared to CBT (Wetherell et al., 2011). ACT is transdiagnostic and associated with improvements in physical functioning and pain-related disability, as well as decreases in emotional distress regardless of perceived pain intensity (Hann & McCracken, 2014).

Resting-state functional magnetic resonance imaging (rsfMRI) allows for data to be collected while individuals with CP rest in the MRI scanner for a short period of time (<10 minutes). Thus, data provides information about the natural state of brain function in CP without having to apply any external sensory or cognitive stimulation. Analysis methods of rsfMRI have focused on multiple regions in the brain, targeting inherent and altered measures of connectivity between brain regions and within brain networks (Fox et al., 2005). Further, alterations in brain structure and function have been demonstrated in multiple CP syndromes (Smallwood et al., 2013, Jensen et al., 2013, Reddan & Wager, 2018). Prior imaging research has suggested that CP results in abnormal *hyper-connectivity* of brain networks associated with self-reflection (default mode, DMN), emotion (salience, SN), and cognitive control (frontal parietal, FPN) networks (Hemington et al., 2016; van Ettinger-Veenstra et al., 2019; Napadow et al., 2010). While ACT has been successful in helping those with CP create a more functional and personally meaningful life (Vowles et al., 2009), a critical gap in our understanding of the neural mechanisms underlying ACT remains.

Only two prior investigations have used fMRI to assess neural mechanisms of ACT-based interventions for CP. Jensen et al. (2012) investigated task fMRI activation using pressure evoked pain. Participants with fibromyalgia showed increased activation in the ventrolateral prefrontal cortex (vlPFC) and orbitofrontal cortex (OFC) post-ACT after 12 weeks of ACT. Additionally, results showed pain-evoked changes in connectivity between the vlPFC and thalamus after ACT. Smallwood and colleagues (2016) conducted an 8-week ACT intervention vs. health education control (HEC) for participants with comorbid CP and opioid addiction. Focusing on DMN and pain regions in the brain, participants receiving ACT exhibited decreased activation during evoked pain in the middle frontal gyrus (MFG), inferior parietal lobule (IPL), insula, anterior cingulate cortex (aCC), posterior cingulate cortex (pCC), and superior temporal gyrus (STG) compared with HEC participants.

In the present study, ACT was delivered to nine women with CP. fMRI was used to identify changes in brain networks underlying ACT-related behavioral outcomes in CP. Based on our prior work examining ACT in CP (Smallwood et al., 2016), we hypothesize that: (1) ACT will reduce connectivity strength within and between the DMN, SN, and FPN, and that (2) changes in connectivity strength will correlate with changes in behavioral outcomes from pre- to post-ACT.

## Methods

### Participants

Nine female participants (47.59 ± 16.54 yrs, 8 right: 1 left-handed) with musculoskeletal pain who did not self-report misusing opioids were enrolled in a 4-week group ACT intervention program (Table 1). Participants were referred from a community-based health care clinic and were required to be at least 18 years of age, speak English, have been living with musculoskeletal CP for 3 or more months, have a Brief Pain Inventory (BPI) Score of >=4, and have no history of cancer or malignancy, head or severe body trauma in the past 6 months. Participants with neurologic (e.g., history of stroke, brain lesions, or intracranial surgery) or psychiatric disorders not commonly comorbid with CP were excluded. Patients who were not addicted to opioids but were taking opiates on a PRN (“as needed”) basis were eligible to participate, in order to reflect real-world clinic conditions as closely as possible. Only one participant self-reported using PRN opioid medication.

**Table 1:** Patient characteristics.

| **Variable** | **Mean** | **Std Dev** | **Minimum** | **Maximum** |
| --- | --- | --- | --- | --- |
| Age | 48 | 18 | 20 | 66 |
| Body Mass Index (BMI) | 24.58 | 3.82 | 20.00 | 32.80 |
| Baseline CES-D Score | 15.22<br>(Median=16) | 6.89 | 4.00 | 25.00 |
| Baseline Pain Severity Score (Brief Pain Inventory (Cleeland & Ryan, 1994)) | 4.17 | 1.57 | 1.75 | 6.25 |
| Years in Pain | 8.7 | 9.2 | 1.5 | 30.0 |

### Acceptance Commitment Therapy Protocol

Patients completed two ninety-minute manualized ACT sessions per week for four weeks (Potter, 2012; Luoma et al., 2007). ACT sessions were administered by two licensed, trained Certified Therapeutic Recreation Specialists (CTRS/L). Behavioral outcomes were measured using validated assessments including the Acceptance & Action Questionnaire (AAQ-II) (Bond et al., 2011), the Center for Epidemiologic Studies Scale (CES-D) (Radloff, 1977; Vilagut et al., 2016), the Chronic Pain Acceptance Questionnaire (CPAQ) (McCracken et al., 2005; 2006; Vowles & Thompson, 2011), the NIH Toolbox *Neuro-QoL™* (Quality of Life in Neurological Disorders) scales (Cella et al., 2012), and the NIH Patient-Reported Outcome Measurement Information System (PROMIS) measures of pain interference (Amtmann et al., 2010; 2011), administered using an iPad. (See supplemental information).

The behavioral assessment data were entered into Excel spreadsheets using Qualtrics software (Qualtrics, 2005) for data management. Statistical analyses were conducted using SAS®v.9.4. (SAS Institute, 2011). Paired Student’s t-tests and Wilcoxon Signed Rank tests were used to assess differences in behavioral measures from pre-to-post ACT (subtracting post minus pre scores). Positive or negative change scores indicated satisfactory results, depending on the specific test in question (e.g. reduced CES-D scores indicated improvements in depression while higher AAQ scores indicated improvements in pain acceptance).

### Resting State fMRI Data Collection

MRI data were collected before and immediately after four weeks of ACT using a Siemens Three Tesla Magnetom Prisma scanner at Boston University, MA. Structural MPRAGE was collected (TR/TE=2.53s/1.32ms, flip angle=7°, field of view (FOV) = 256 × 320 mm, 0.8mm^3^ resolution) to allow for anatomical registration. Subsequently, two runs of eight minutes resting-state fMRI data were obtained using a T2^*^ weighted Echo Planar Imaging sequence (2.5mm^3^ resolution, 60 slices, TR/TE=1.2s/30ms, 300 volumes, FOV=205mm, multi-slice interleaved ascending) for all participants. During the resting state scans, participants were instructed to lie still in the scanner with their eyes open, fixating on a crosshair placed in their field of view. Only the first of the two resting state scans were used for analysis. Two rsfMRI scans were collected in the case that one set was unusable (e.g. movement artifact). The first scan set was of high enough quality to use.

### Resting State fMRI Data Analysis

Standard preprocessing steps were carried out using Statistical Parametric Mapping, version 12 (SPM12, Penny et al., 2006). First, all scan data were imported in the form of DICOM images and converted to Nifti files using the DICOM Import function in SPM12. Functional data were realigned and co-registered to the standard Montreal Neurological Institute (MNI) template in SPM12. Motion correction, band-pass filtering (0.0078 – 0.08 Hz), slice-timing correction, normalization to remove individual variability for between subject comparisons, and smoothing to increase signal to noise ratio were carried out using SPM12 (Figure 1, step 1). Next, each participant’s brain was parcellated into discrete regions of interest representing nodes from the Power atlas (Power et al., 2011) using Mango (Multi-image Analysis GUI; Lancaster et al., 2010). The mean time course within seed regions were extracted from the residual images using REX (Duff, 2008; Figure 1, step 2). FC estimates were then calculated using the pairwise Pearson correlation of seed regions located time course across all 264 nodes resulting with a 264 by 264 connectivity matrix (Figure 1, step 3). Finally, connectivity matrices were reduced to a subset of 101 nodes that were associated with the FPN, DMN, and SN.

**Figure 1:**
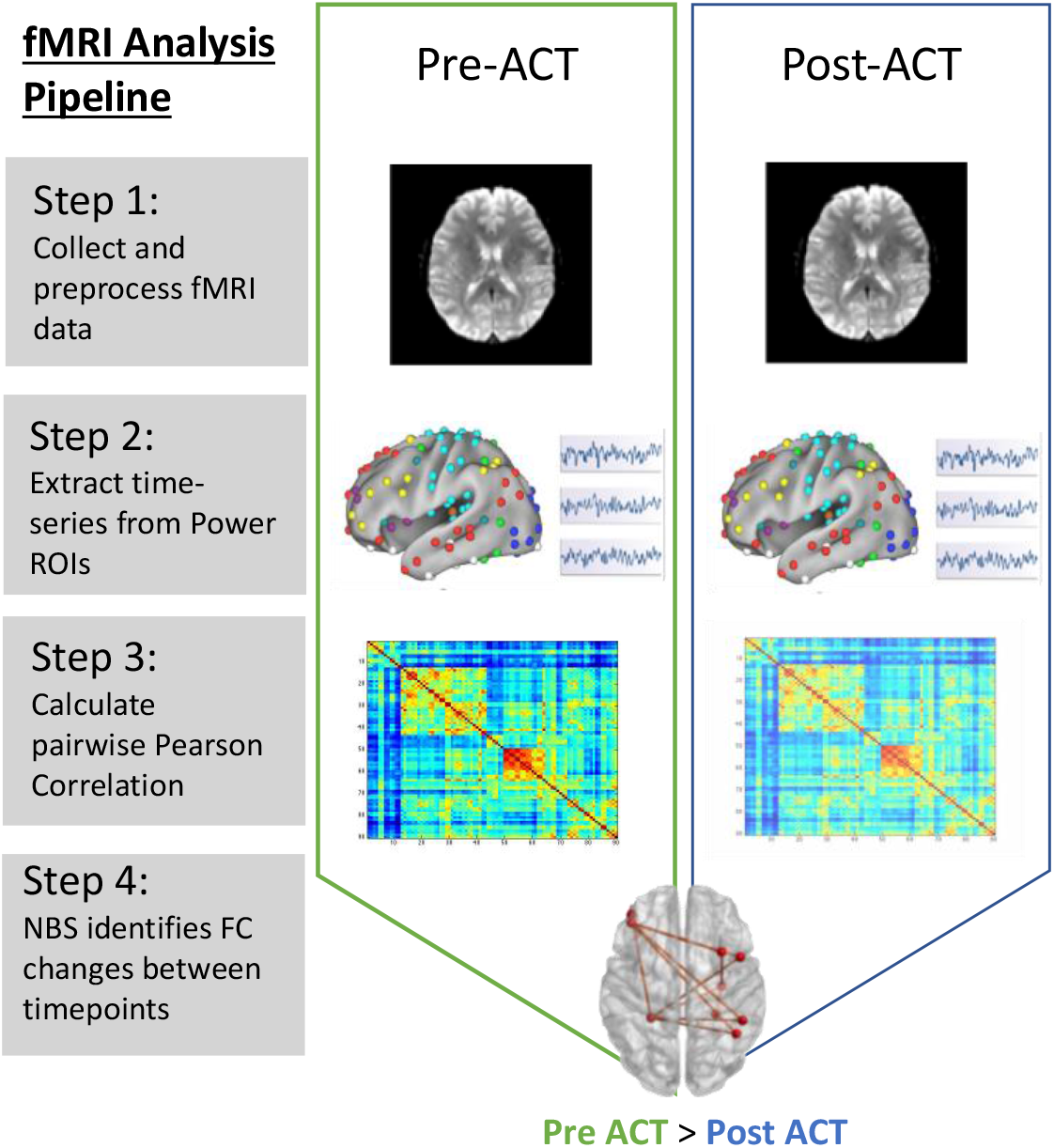
fMRI data analysis pipeline. Step 1: rsfMRI data preprocessed using SPM12; Step 2: Mean time series extraction from Power ROIs; Step 3: Estimate FC between all nodes using Pearson correlation; Step 4: Use NBS to identify changes in connectivity between pre- and post-ACT.

### Graph Analysis

Graph analysis applied to fMRI is a powerful way of characterizing brain networks. In this context, a network represents a collection of nodes, and the functional connections between pairs of nodes. Nodes in large-scale brain networks represent brain regions, with connections being anatomical, functional, or effective, depending on the type of imaging data analyzed. Application of graph theory-based approaches have identified biologically plausible brain networks found to topologically organize in a non-trivial manner (e.g., network integration and modular structure) and support efficient information processing of the brain (Wang et al., 2010, Bullmore et al., 2009; 2011). These network analyses allow us to visualize the connectivity pattern across the entire brain and also quantitatively characterize its global organization (Wang et al., 2010; Sporns, 2018). In the current study, we leverage the ability of graph analytics to identify network connections linking rsfMRI connectivity to ACT treatment.

### Network Based Statistic

We used the Network Based Statistic (NBS; Zalesky et al., 2010) to examine FC in the DMN, SN, and FPN. This graph theory-based method provides a statistical approach to identify changes in FC associated with diagnostic status or changing psychological contexts (Zalesky et al., 2010). The NBS is based on the principles underpinning traditional cluster-based thresholding of statistical parametric maps. We use the NBS to identify significant network connectivity differences between pre-ACT and post-ACT (Figure 1, step 4). We tested for within network connectivity changes for each of the 3 networks of interest independently, and then we tested for network connectivity changes across all 101 nodes that make up the FPN, DMN, and SN. Results presented represent functional network differences for t > 2.5 (10,000 permutations). We further examine whether specific pairwise connections in brain networks affected by ACT are related to behavior, and whether connectivity changes associated with ACT correlate with changes in behavior/outcome measures (using Pearson’s *r*).

### Causal Mediation Analysis

Causal mediation analyses were conducted, with confounder adjustment, to evaluate associations between FC changes and behavioral changes (pre- to post-ACT). In other words, we tested whether connectivity changes reflected by the NBS (Figure 1, step 4) were mediators of changes in depression, social role, and CP acceptance scores. A counterfactual mediation approach was implemented in this analysis using SAS PROC CAUSALMED (Valeri & VanderWeele, 2013; VanderWeele, 2015). Although structural equation modeling (SEM) is frequently used to examine mediation (Baron & Kenny, 1986), its limitations include the following: (1) It does not have a clear theoretical foundation for defining causal mediation effects; (2) It does not deal with confounding and interaction effects effectively; (3) It does not treat binary outcomes and binary mediators in a unified manner. The counterfactual approach overcomes these limitations (SAS Institute, 2019; MacKinnon et al., 2007).

Causal mediation analysis quantifies and estimates the total, direct, and indirect (or mediated) effects between an independent variable and an outcome. It enables causal interpretations of these effects under the assumptions of the counterfactual framework (Robins & Greenland, 1992; Pearl, 2011). The causal mediation model decomposes the total effect into a direct effect (e.g., the effect of an independent variable [A] on outcome [Y; A = 0 vs. A^*^=1]) and the natural indirect effect (NIE)). The controlled direct effect (CDE) simulates a randomized controlled trial (RCT) by hypothetically assigning the same value of the mediator to all individuals in the sample, with bootstrapped standard errors (Naimi et al., 2014). The NIE captures the effect of the mediation pathway (e.g., the average change in Y if the exposure is fixed to the level of the intervention and the mediator changes accordingly [e.g., A = 0 to A^*^ =1]; VanderWeele, 2015). The mediation path is represented by arrows “B” and “C” in Figure 2.

**Figure 2:**
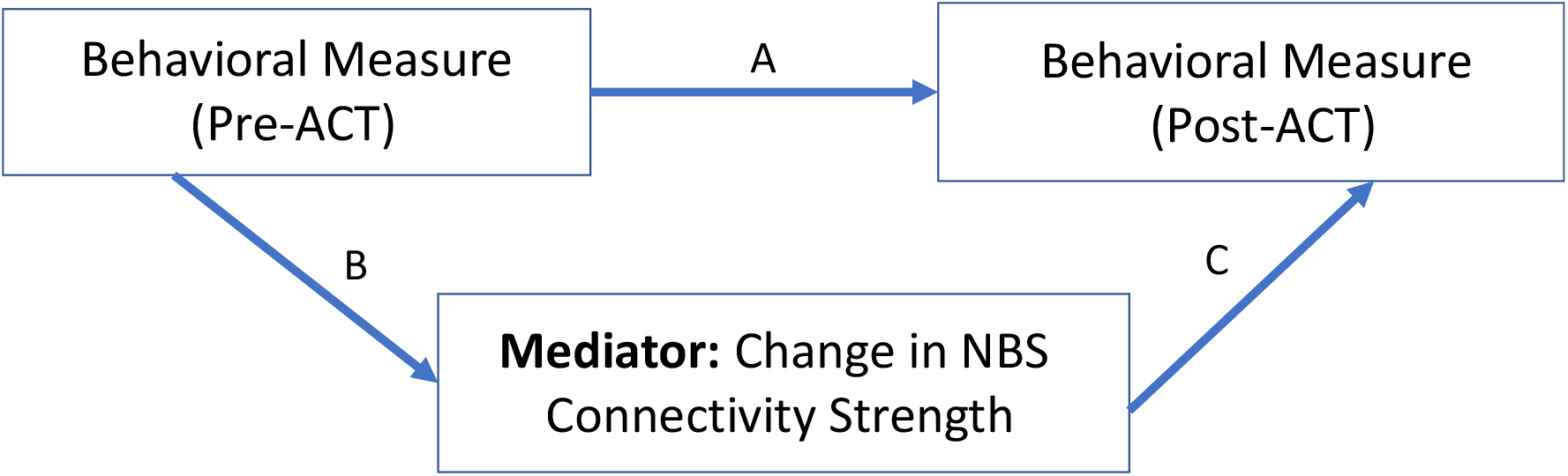
Rationale for the causal mediation analysis. Change in behavioral measures (pre- to post-ACT), mediated by FC changes. Path “A” shows the direct effect; mediation is estimated by combining paths B and C to produce the NIE.

For this analysis, we used the theoretical constructs underlying ACT to guide our approach (e.g., that *psychological flexibility*, in the context of CP, includes factors pertaining to acceptance and cognitive defusion (learning to experience uncomfortable thoughts, feelings, and sensations in a way that reduces their interference on valued activities and roles in one’s daily life); Chin & Hayes, 2017). Thus, we focused on indicators of CP acceptance, pain interference, depression, and social roles as key behavioral outcomes.

## Results

Behavioral change scores from pre-to post-ACT showed statistically significant improvements in clinically relevant outcomes, including depression (measured via the CES-D and the NIH Toolbox Neuro-QoL^TM^), satisfaction with social role (measured via the NIH Toolbox Neuro-QoL^TM^), and pain acceptance (measured via the AAQ-II and CPAQ) (Table 2). Two specific sub-scores of the CPAQ indicated that there were significant decreases in pain interference and significant increases in willingness to engage in activities despite pain.

**Table 2:** Behavioral Change Measures. Descriptive statistics and changes in behavioral outcome scores from pre- to post-ACT.

| Variable | Pre-ACT Mean(StDev) | Post-ACT Mean(StDev) | Change Post-Pre ACT Mean(StDev) | Pr > \|t\| (Paired t-Test) | Non-Parametric Pr>= \|S\| (Wilcoxon Signed Rank Test) |
| --- | --- | --- | --- | --- | --- |
| Social Roles (NIH Toolbox) | 20(5.87) | 28.78(6.46) | 8.78(6.51) | 0.0037 | 0.0039 |
| Depression (NIH Toolbox) | 12.11(5.49) | 8.67(4.03) | -3.44(3.28) | 0.0136 | 0.0313 |
| CES-D | 15.22(6.89) | 9.11(5.35) | -6.11(5.84) | 0.0138 | 0.0273 |
| AAQ-II | 47.44(7.26) | 54.33(8.8) | 6.89(7.25) | 0.0215 | 0.0156 |
| CPAQ (total) | 71.22(13.89) | 84.44(14.68) | 13.22(14.02) | 0.0222 | 0.0156 |
| Pain Willingness (CPAQ) | 34(6.44) | 39.11(4.37) | 5.11(5.44) | 0.0226 | 0.0156 |
| Pain Interference (PROMIS) | 3.86 (1.65) | 1.80 (1.48) | -1.86 (1.54) | 0.0113 | 0.0234 |
| Pain Severity (BPI) | 4.17(1.57) | 3.69(1.95) | -0.41(1.32) | 0.4137 | 0.5625 |

### Network Based Statistic

Significant changes in FC from pre- to post-ACT in the DMN, SN, and FPN were observed. Using the NBS, *within* network effects of ACT were only observed in the SN which consisted of a sub-network of four nodes and three functional connections (Figure 3A, *t*>2.5, *p*=0.039). No effects of ACT were observed within the DMN or FPN. NBS tests comparing preACT vs post-ACT of the triple network (DMN, FPN, and SN nodes combined) identified a network of 10 nodes and 10 connections displaying decreases in FC associated with completing ACT (Figure 3B, *t*>2.5, *p*=0.05). Interestingly, the within network ACT effects observed in the SN were also present in the triple network. Between network changes were also observed in the triple network where all DMN and FPN nodes connected to SN nodes (Figure 3B).

**Figure 3:**
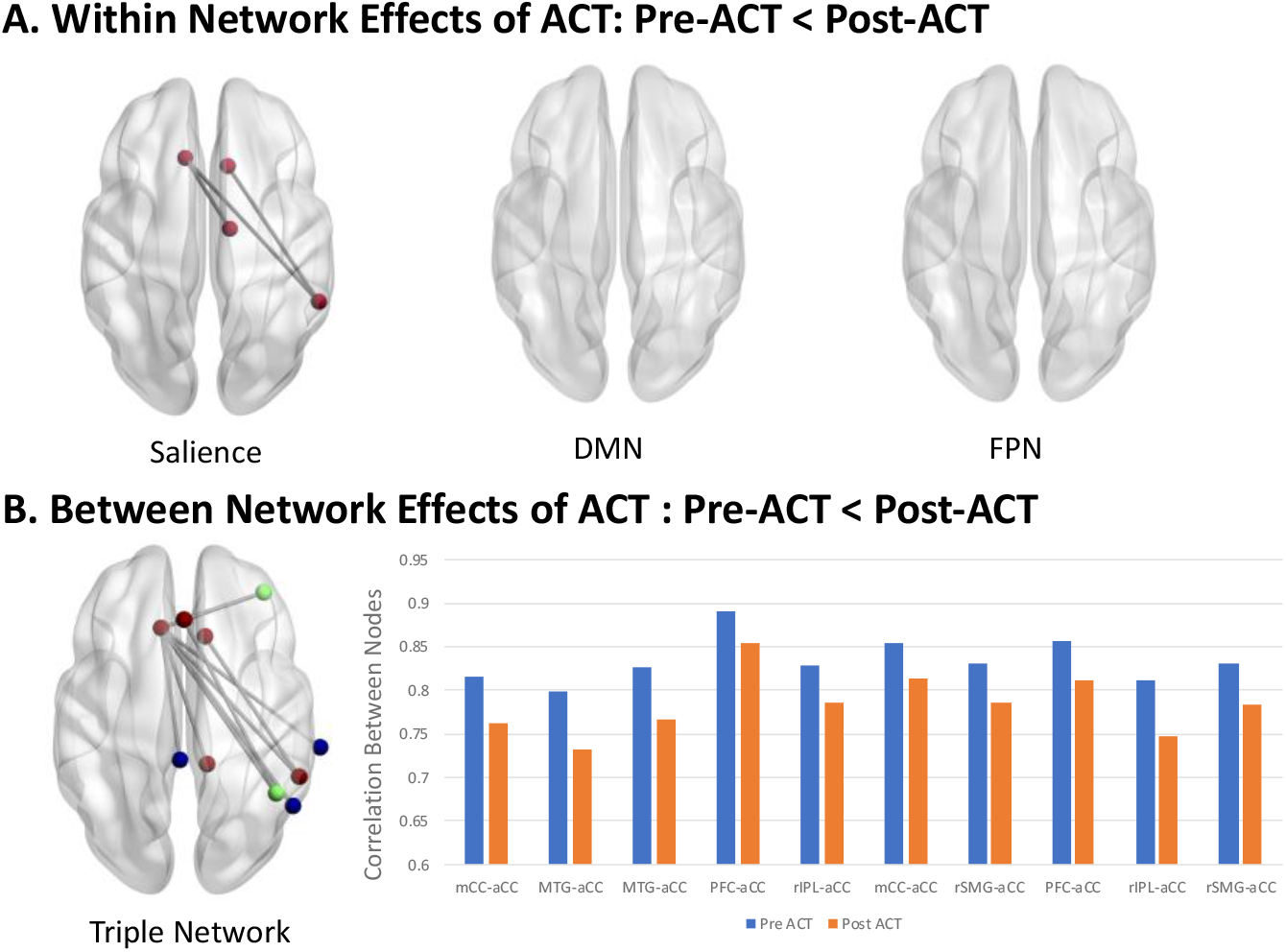
Within and between network effects of ACT. A) Within network decreases in FC following ACT: Pre-ACT < Post-ACT (t>2.5, p =0.039, 10,000 iterations). B) Between network decreases in FC following ACT: Pre-ACT < Post-ACT (t>2.5, p =0.05, 10,000 iterations). Mean functional connections exhibiting ACT effects are shown for Pre-ACT (blue) and Post-ACT (orange). SN, DMN, and FPN nodes are shown in red, blue, and green respectively. mCC = midcingulate cortex, aCC = anterior cingulate cortex, MTG = medial temporal gyrus, PFC = prefrontal cortex, rIPL = right inferior parietal cortex, rSMG = right superior medial gyrus. Node numbering below anatomical labels correspond to the node ordering in the Power Atlas. (See supplemental information).

### Correlation Analysis

Next, we assessed the relationships between brain connectivity changes and behavioral outcomes using Pearson correlation statistics. Six of the ten functional connections that showed significant differences in strength within the Triple Network from pre- to post-ACT (shown in Figure 3B) were significantly correlated with behavior change scores. Pearson correlations and corresponding p-values for the following significant relationships are shown in Table 3. Results indicated that ACT effects in connectivity (functional connections from NBS analyses shown in Figure 3) between the MTG and aCC (52, −59, 36; −11, 26, 25) and between the PFC and aCC (38, 43, 15; 0, 30, 27) were correlated with social role (*Neuro-QoL*^TM^) and acceptance (AAQ-II) scores. FC changes between the rIPL and aCC (two functional connections connecting 44, −53, 47 with −11, 26, 25 and 0, 30, 27) were correlated with *Neuro-QoL™* depression scores; and changes between rSMG and aCC (55, −45, 37; 10, 22, 27) were correlated with both depression and social role scores. Changes between rSMG and aCC (55, - 45, 37; −11, 26, 25) and rIPL and aCC (44, −53, 47; −11, 26, 25) were correlated with reduced pain severity scores.

**Table 3:** Correlations between functional connections in Triple network and behavioral assessment scores (post minus pre). Pearson’s *r* is value shown in top half of each cell, its corresponding p-value is shown in the bottom half of each cell. Significant values are indicated with bold font (*p=0.05 (**meets Bonferroni corrected p=0.005)).

| Functional Connection | Depression Change Score<br>*r*<br>(*p*) | CES-D Change Score<br>*r*<br>(*p*) | Social Role Change Score<br>*r*<br>(*p*) | AAQ-II Change Score<br>*r*<br>(*p*) | CPAQ Pain Willingness Change Score<br>*r*<br>(*p*) | Pain Severity Change Score<br>*r*<br>(*p*) |
| --- | --- | --- | --- | --- | --- | --- |
| MTG-aCC | -0.3621 | -0.2123 | **0.8618** | **0.8681** | 0.4225 | -0.5462 |
|  | 0.3383 | 0.5834 | **0.0028\*\*** | **0.0024\*\*** | 0.2572 | 0.1614 |
| PFC-aCC | -0.2107 | 0.1491 | **0.6820** | 0.4767 | -0.2353 | -0.1293 |
|  | 0.5863 | 0.7018 | **0.0430\*** | 0.1945 | 0.5423 | 0.7602 |
| rIPC-aCC | **-0.6848** | **-0.6881** | 0.6266 | 0.4254 | **0.7268** | **-0.8415** |
|  | **0.0418\*** | **0.0405\*** | 0.0709 | 0.2536 | **0.0265\*** | **0.0088\*** |
| rIPC-aCC | **-0.7432** | -0.3970 | 0.5408 | 0.2222 | 0.2092 | -0.5112 |
|  | **0.0218\*** | 0.2901 | 0.1328 | 0.5656 | 0.5890 | 0.1954 |
| rSMG-aCC | **-0.7290** | -0.1363 | **0.7001** | 0.5143 | 0.1058 | -0.1995 |
|  | **0.0259\*** | 0.7266 | **0.0357\*** | 0.1566 | 0.7865 | 0.6357 |
| rSMG-aCC | -0.41367 | -0.65678 | 0.44638 | 0.30597 | 0.6166 | **-0.8668** |
|  | 0.2684 | 0.0546 | 0.2284 | 0.4233 | 0.0770 | **0.005\*\*** |

### Causal Mediation Model Results

In the causal mediation framework for this study, a baseline behavioral variable (e.g., Pre-ACT Depression Score) was hypothesized to relate to an outcome variable (Post-ACT Depression score) via the causal mechanism that is represented in Figure 2. As depicted in the diagram, the total causal treatment effect pertaining to the outcome (Post-treatment behavioral score) consists of the following two parts: (1) a direct effect; (2) a mediated (or indirect) effect via the “functional connectivity” variable representing change in FC between two brain regions.

Results from our exploratory causal mediation models (Table 4) demonstrated that improvements in specific behavioral outcome scores were significantly related to both the direct pathway “A” (e.g., baseline behavior scores predicted post-treatment scores) and through the indirect mediation pathway “B and C” via changes in FC (the NIE). Statistically significant NIE provide evidence in support of our mediation hypothesis for rSMG-aCC for depression, MTG-aCC for social role, both rSMG-aCC and MTG-aCC for CP acceptance, and rlPL-aCC for pain interference. These relationships persisted after covariate adjustment for age, BMI, pain severity, and other potential confounders. By contrast, statistically significant mediation effects were not observed for rlPL-aCC for depression, PFC-aCC and social role, or MTG-aCC and AAQ-II. No significant interactions were detected.

**Table 4:** Causal Mediation Model Results. Relationships between hypothesized functional connectivity mediators and behavioral outcome scores (post-minus pre-ACT). Functional connectivity mediator is measured as the change (post-minus pre-ACT) in the NBS. Independent variable = Pre-ACT Behavioral Score; Dependent variable= Post-ACT Behavioral Score. Controlled Direct Effect (CDE) simulate an RCT by hypothetically assigning the same value of the mediator to the individuals in the sample (Naimi et al., 2014).

| Hypothesized Functional Connectivity Mediator<sup>1</sup> | Behavioral Measure<sup>2</sup> | Coefficient ($\beta$) for Controlled Direct Effect Estimate (CDE)<sup>3</sup><br>(Bootstrap 95% Confidence Intervals) | p-value Direct for Effect (path A) | Mediation: Coefficient ($\beta$) for Natural Indirect Effect Estimate (NIE)<br>(Bootstrap 95% Confidence Intervals) | p-value for Mediation (paths B, C) | Covariates (Control factors) |
| --- | --- | --- | --- | --- | --- | --- |
| rSMG-aCC | Depression (NeuroQOL) | 0.986<br>(0.89, 1.082) | <0.0001 | -0.231<br>(-0.372, -0.091) | 0.0012 | Age, BMI, Pain Severity, Baseline Social Role Score |
| rIPL-aCC | Depression (NeuroQOL) | 0.721<br>(0.657, 0.786) | <0.0001 | 0.033<br>(-0.81, 0.148) | 0.5686 | Age, BMI, Pain Severity, Baseline Social Role Score |
| MTG-aCC | Social Role (NeuroQOL) | 1.645<br>(2.150, 1.049) | <.0001 | -1.733<br>(-2.428, -1.038) | <.0001 | Age, BMI, Pain Severity, Baseline Depression Score |
| PFC-aCC | Social Role (NeuroQOL) | 0.292<br>(-0.011, 0.595) | 0.0591 | 0.154<br>(-0.459, 0.815) | 0.6063 | Age, BMI, Pain Severity, Baseline Depression Score |
| MTG-aCC | Acceptance & Action (AAQ-II) | 1.395<br>(1.195, 1.50) | <.0001 | -0.062<br>(-0.197, 0.072) | 0.3620 | Age, BMI, Baseline Depression Score; Pain Severity |
| rSMG-aCC | Chronic Pain Acceptance (CPAQ total) | -1.088<br>(-1.144, -1.031) | <.0001 | 1.060<br>(0.813, 1.306) | <.0001 | Age, BMI, Pain Severity, Baseline Depression Score, Baseline Social Role Score |
| MTG-aCC | Chronic Pain Acceptance (CPAQ Total) | -0.452<br>(-0.675, -0.228) | <.0001 | 0.424<br>(0.153, 0.695) | 0.0022 | Age, BMI, Pain Severity, Baseline Depression Score, Baseline Social Role Score |
| rIPL-aCC | CPAQ - Pain Willingness | -0.020<br>(0.029, -0.009) | 0.0001 | 0.002<br>(-0.029, -0.010) | 0.6091 | Age, BMI, Pain Severity, Baseline Depression Score, Baseline Social Role Score |
| rIPL-aCC | Pain Interference (PROMIS) | 1.970<br>(1.057, 2.882) | <.0001 | -0.1679<br>(-2.870, -0.529) | 0.0044 | Age, BMI, Baseline Depression Score, Baseline Social Role Score |
| rSMG-aCC | Pain Severity | 0.810<br>(0.474, 1.146) | <.0001 | -0.507<br>(0-.950, -.065) | 0.0247 | Age, BMI, Baseline Depression Score, Baseline Social Role Score |
| rIPL-aCC | Pain Severity | 1.2756<br>(1.225, 1.326) | <.0001 | -0.9725<br>(-1.426, -0.519) | <.0001 | Age, BMI, Baseline Depression Score, Baseline Social Role Score |

## Discussion

We examined rsfMRI data of nine women before and after completing ACT in efforts to better understand changes in brain FC and associations with specific behavioral outcomes. Importantly, we used NBS to assess network function with graph analyses and took an innovative approach to study of the relationship between imaging and behavioral measures known as causal mediation analysis. Our results showed significant improvements from pre-to post-ACT in clinically relevant behavioral outcomes, including depression, satisfaction with social role, and pain acceptance. These results confirm findings from other studies (McCracken et al., 2004, 2005, 2006; Wetherell et al., 2011; 2016) and align with the theoretical principles underlying ACT, specifically that constructs of psychological flexibility including acceptance and cognitive defusion (reduced pain interference) may play important roles in functional improvements for individuals suffering from CP (Chin & Hayes, 2017). ACT does not aim to limit exposure to negative experiences, but encourages persons with CP to decrease attention to pain and to move forward in valued life directions despite these experiences. In our study, two specific sub-scores of the CPAQ indicated that there were significant decreases in pain interference and significant increases in willingness to engage in activities despite pain. Other researchers studying mind-body therapies have documented similar results. For example, Haugmark et al. (2019) analyzed health effects of mindfulness- and acceptance-based interventions, including mindfulness-based stress reduction (MBSR), mindfulness-based cognitive therapy (MBCT), and ACT. The authors found small to moderate effects in favor of mindfulness- and acceptance-based interventions compared to controls in pain, depression, anxiety, mindfulness, sleep quality, and health-related quality of life.

We used the NBS (Zalesky et al., 2010) to examine FC in the DMN, SN, and FPN and found within SN effects that extend to brain regions that are components of the DMN and FPN. Prior fMRI studies on mindfulness approaches for treating CP have shown that increased regional activation in the aCC and OFC were associated with reduced ratings of the unpleasantness of pain (Zeidan et al., 2011; 2012; 2015; Lazaridou et al., 2016, Yoshino et al., 2018). Additional multidisciplinary pain treatment programs comprised of daily physical and occupational therapies plus CBT for pain treatment resulted in improved pain measures that correlated with connectivity changes in DMN, SN, and FPN (Simons et al., 2014). Specifically, findings showed treatment driven reductions of hyper-connectivity from the left amygdala to the motor cortex, parietal lobe, and CC. Simons et al. (2014) also found that connectivity to several regions of the fear circuitry (PFC), bilateral middle temporal lobe, bilateral CC, and hippocampus correlated with higher pain-related fear scores, and that decreases in pain-related fear correlated with decreased connectivity among the amygdala and the motor and somatosensory cortex, CC, and the FPN. Across the few studies utilizing longitudinal randomized controlled designs with active control groups, aCC, PFC, pCC, insula, striatum (caudate, putamen), and amygdala show relatively consistent changes associated with mindfulness meditation (Tang et al., 2012; 2016; Hölzel et al., 2011).

These studies, in conjunction with our results, suggest that the neural correlates of ACT for CP affect sensory brain networks and cognitive function. Thus, our results suggest that the neural mechanisms underlying the multi-faceted nature of ACT for CP are not only related to the DMN, but also to the DMN’s relationship to other networks. A consistent finding across several studies is that CP results in hyper-connectivity among the DMN, SN (Hemington et al., 2016; van Ettinger-Veenstra et al., 2019), and FPN (Napadow et al., 2010). Supporting the current analyses, the transition from acute to CP over a 1-year period has been found to result in a gradual ‘shift’ in fMRI activations from nociceptive networks to emotional brain networks (Hashmi et al., 2013). Collectively, these findings provide impetus for further study of associations between rsfMRI and clinical outcomes.

### Behavioral Outcomes and fMRI

Several studies have evaluated the association between fMRI and behavioral outcomes in mindfulness-based interventions (e.g., MBSR), though the majority of these evaluations are not specific to ACT (Tang & Leve, 2016; Zeidan et al., 2018, Feliu-Soler et al., 2018). A recent study (Yoshino et al., 2018) used a CBT intervention with behavioral activation (an ACT component) and found that the OFC played an important role in improvements in pain intensity post-treatment. Activation of the dorsal pCC at pre-treatment was also associated with improvements of clinical symptoms. ACT has also demonstrated sustained medium-large effect sizes on social functioning (Dahl, Wilson, & Nilsson, 2004; Feliu-Soler et al., 2018).

Yu et al., (2020) examined ACT-oriented treatment for fatigue in 354 adults with CP. Pearson’s correlations and hierarchical regression were conducted to investigate associations between improvement in fatigue interference and improvements in outcomes. Mixed effects models were used to explore associations between baseline fatigue interference and changes in outcome measures. Results suggested that participants improved in perceptions of fatigue interference, pain, Psychological Flexibility (PF) processes, and daily functioning. Changes in fatigue interference were associated with changes in pain, PF processes, and daily functioning |r| = 0.20−0.46. Changes in fatigue interference were associated with changes in pain acceptance independent of changes in pain perception. The authors concluded that individuals with fatigue appeared to benefit from the ACT-oriented interdisciplinary treatment for CP, and relatively higher levels of fatigue did not appear to decrease the treatment benefit. Although fatigue was not a focus in our study, the results are similar in terms of demonstrating that the behavioral improvements associated with ACT may persist regardless of pain severity and the presence of other co-morbidities.

Smallwood et al. (2013) examined gray matter volume (GMV) differences between CP patients and healthy controls and found that there were 12 clusters where GMV was decreased in CP patients compared with controls. These clusters included many regions that are considered part of the “pain matrix” involved in pain perception, but also included many other regions that are not commonly regarded as pain-processing areas. The authors also reported that the most common behavioral domains associated with these regions were cognitive, affective, and perceptual domains, suggesting that many of the regions may relate to the constellation of comorbidities that often accompany CP (e.g., fatigue, depression, cognitive, and emotional impairments).

### Integrating Behavioral and Neural Network Changes using Causal Mediation Analysis

Using causal mediation analysis to assess whether changes in connectivity strength mediated changes in specific behavioral outcomes, we observed statistically significant mediation effects for rSMG-aCC with depression, MTG-aCC with social role, rSMG-aCC and MTG-aCC with CP acceptance, and rlPL-aCC with pain interference. Because the models were adjusted for age, BMI, pain severity, and other behavioral covariates, we were able to determine that the relationships were not confounded by these factors.

We also observed significant mediation effects for rSMG-aCC and rlPL-aCC with pain severity, despite the fact that changes in perceived pain severity are not considered direct targets of ACT. In our unadjusted analyses, perceived pain severity scores did not change significantly from pre- to post-ACT, yet the controlled direct effect in the causal mediation models, with confounder adjustment, demonstrated significant changes in both direct and indirect effects. These exploratory analyses underscore the complexity of measuring pain perception, particularly as other therapeutically targeted behavioral changes and associated neural connectivity changes may be occurring simultaneously.

Notably, the median baseline CES-D score among our participants was 16, indicative of high depressive symptomatology (Vilagut et al., 2016) concurrent with CP. After the ACT intervention, the median CES-D score was reduced to 7, and this change appears to be mediated by decreased rSMG-aCC hyperconnectivity.

By contrast, statistically significant mediation effects were not observed for rlPL-aCC with depression, MTG-aCC with AAQ-II, or PFC-aCC with social role. In these cases, the relationships were confounded by other factors and may operate via more complex multiple mediation pathways that could not be tested in this small exploratory sample. For example, although we observed a statistically significant mediation effect for PFC-aCC and social role in unadjusted models, inclusion of body mass index (BMI), pain severity, and baseline depression nullified this relationship.

A growing body of literature has begun to employ mediation analysis to assess the mechanisms underlying behavioral and clinical outcomes (Atlas et al., 2014; Cederberg et al., 2016; DasMahapatra et al., 2015; Gu et al., 2015; Lee et al., 2015; Mischkowski et al., 2019; Sanders et al., 2017; Whibley et al., 2019). However, few studies have assessed behavioral changes with respect to fMRI data using causal mediation analysis, particularly with respect to CP. Lindquist (2012) described an extension of SEMs applied to data from a fMRI study of thermally induced pain. The results suggested that many classic “pain-responsive regions” such as the anterior insula showed significant mediation of the temperature-induced relationship, and that subjective pain ratings increased near the end of the stimulation period. Other regions, such as the insular cortex appeared to be active during pain judgment. Like our study, this study supports the use of mediation modeling in future research to better understand how connectivity changes among different brain regions affect the subjective experience of pain, and may inform pain management approaches.

## Limitations and Future Directions

In addition to the small sample size, the limitations of our study include lack of a randomized control group and lack of long-term follow-up data beyond the immediate post-ACT period.

Villaneuva et al. (2019) investigated three key aspects of ACT, including its effectiveness, long-term follow-up, social context, and social processes. The authors contend that researchers should include longer follow up periods in clinical studies (Gloster et al., 2013). This is especially important for treatment-resistant patients (e.g., patients who do not respond to standard, first line treatments), for whom viable treatment options are limited (Gloster et al., 2015).

Factors outside therapy itself, including social processes, may account for up to 33% of improvement in patients undergoing psychotherapy and group-based interventions (Cuijpers et al., 2012). It remains poorly understood how the influence of social surroundings longitudinally affects patients’ well-being, social function, and pain perception (Biglan & Embry, 2013; Wilson et al., 2014). Prior research suggests that both close and extended social ties may be relevant for positive outcomes (Kuehner & Huffziger, 2013). Additional research is needed to better understand the variation in treatment outcomes in relation to an individual’s social and environmental context, using an exposome lens (Juarez et al., 2014). Future research should also consider different ways of measuring pain perception and should evaluate both mediators and moderators of ACT in pain as well as in other chronic diseases.

## Conclusions

The mechanistic knowledge generated from this study helps to build the evidence base underlying mind-body therapies such as ACT. ACT has been shown to be particularly efficacious for patients who are older (Wetherell et al., 2016) or have co-occurring mood disorders (Wolitzky-Taylor et al., 2012) who may be unresponsive to other psychosocial treatments. Findings from the present study facilitate identification of neural factors predicting patient responses to mind-body therapies. The outcomes of this study will also support the refinement of non-pharmacologic treatment protocols for CP. This is particularly important with the movement towards ‘stepped care’ models of pain management (Blair et al., 2015), which aim to treat pain within a primary care setting while enabling the use of a variety of integrated multidisciplinary treatment approaches.

## Data Availability

Data is provided in Supplemental Information; please contact the authors for other data questions.

## Acknowledgements

We thank Tye Thompson, Airi Mooers, Bronwyn Leto, Grace Roy, Jennifer Potter, Kristen Rosen, Kathryn Kanzler, Robert Ross, Kerry Nolte, Linda Mamakos, Joseph Haviland, Julia Bushell, Evelyn DeRosa, and Jessica Davis for their assistance with this study.

## Supplemental Information

**Table 1:** Power et al. 2011 nodes were used in graph analyses. Listed below are ROIs corresponding to the DMN, FPN, and SN:

| Power ROI | X | Y | Z | Network | Region |
| --- | --- | --- | --- | --- | --- |
| 74 | -41 | -75 | 26 | Default mode | Occipital_Mid_L |
| 75 | 6 | 67 | -4 | Default mode | Frontal_Mid_Orb_R |
| 76 | 8 | 48 | -15 | Default mode | Frontal_Mid_Orb_R |
| 77 | -13 | -40 | 1 | Default mode | Lingual_L |
| 78 | -18 | 63 | -9 | Default mode | Frontal_Sup_Orb_L |
| 79 | -46 | -61 | 21 | Default mode | Temporal_Mid_L |
| 80 | 43 | -72 | 28 | Default mode | Occipital_Mid_R |
| 81 | -44 | 12 | -34 | Default mode | Temporal_Pole_Mid_L |
| 82 | 46 | 16 | -30 | Default mode |  |
| 83 | -68 | -23 | -16 | Default mode |  |
| 86 | -44 | -65 | 35 | Default mode | Angular_L |
| 87 | -39 | -75 | 44 | Default mode | Angular_L |
| 88 | -7 | -55 | 27 | Default mode | Precuneus_L |
| 89 | 6 | -59 | 35 | Default mode | Precuneus_R |
| 90 | -11 | -56 | 16 | Default mode | Precuneus_L |
| 91 | -3 | -49 | 13 | Default mode | Precuneus_L |
| 92 | 8 | -48 | 31 | Default mode | Cingulum_Post_R |
| 93 | 15 | -63 | 26 | Default mode | Precuneus_R |
| 94 | -2 | -37 | 44 | Default mode | Cingulum_Mid_L |
| 95 | 11 | -54 | 17 | Default mode | Precuneus_R |
| 96 | 52 | -59 | 36 | Default mode | Angular_R |
| 97 | 23 | 33 | 48 | Default mode | Frontal_Sup_R |
| 98 | -10 | 39 | 52 | Default mode | Frontal_Sup_Medial_L |
| 99 | -16 | 29 | 53 | Default mode | Frontal_Sup_L |
| 100 | -35 | 20 | 51 | Default mode | Frontal_Mid_L |
| 101 | 22 | 39 | 39 | Default mode | Frontal_Sup_R |
| 102 | 13 | 55 | 38 | Default mode | Frontal_Sup_Medial_R |
| 103 | -10 | 55 | 39 | Default mode | Frontal_Sup_L |
| 104 | -20 | 45 | 39 | Default mode | Frontal_Sup_L |
| 105 | 6 | 54 | 16 | Default mode | Frontal_Sup_Medial_R |
| 106 | 6 | 64 | 22 | Default mode | Frontal_Sup_Medial_R |
| 107 | -7 | 51 | -1 | Default mode | Cingulum_Ant_L |
| 108 | 9 | 54 | 3 | Default mode | Frontal_Sup_Medial_R |
| 109 | -3 | 44 | -9 | Default mode | Frontal_Mid_Orb_L |
| 110 | 8 | 42 | -5 | Default mode | Frontal_Mid_Orb_R |
| 111 | -11 | 45 | 8 | Default mode | Cingulum_Ant_L |
| 112 | -2 | 38 | 36 | Default mode | Frontal_Sup_Medial_L |
| 113 | -3 | 42 | 16 | Default mode | Cingulum_Ant_L |
| 114 | -20 | 64 | 19 | Default mode | Frontal_Sup_L |
| 115 | -8 | 48 | 23 | Default mode | Frontal_Sup_Medial_L |
| 116 | 65 | -12 | -19 | Default mode |  |
| 117 | -56 | -13 | -10 | Default mode | Temporal_Mid_L |
| 118 | -58 | -30 | -4 | Default mode | Temporal_Mid_L |
| 119 | 65 | -31 | -9 | Default mode | Temporal_Mid_R |
| 120 | -68 | -41 | -5 | Default mode |  |
| 121 | 13 | 30 | 59 | Default mode |  |
| 122 | 12 | 36 | 20 | Default mode | Cingulum_Ant_R |
| 123 | 52 | -2 | -16 | Default mode | Temporal_Mid_R |
| 124 | -26 | -40 | -8 | Default mode | ParaHippocampal_L |
| 125 | 27 | -37 | -13 | Default mode | ParaHippocampal_R |
| 126 | -34 | -38 | -16 | Default mode | Fusiform_L |
| 127 | 28 | -77 | -32 | Default mode | Cerebelum_Crus1_R |
| 128 | 52 | 7 | -30 | Default mode | Temporal_Mid_R |
| 129 | -53 | 3 | -27 | Default mode | Temporal_Mid_L |
| 130 | 47 | -50 | 29 | Default mode | Angular_R |
| 131 | -49 | -42 | 1 | Default mode | Temporal_Mid_L |
| 137 | -46 | 31 | -13 | Default mode | Frontal_Inf_Orb_L |
| 139 | 49 | 35 | -12 | Default mode | Frontal_Inf_Orb_R |
| **174** | -44 | 2 | 46 | Fronto-parietal Task Control | Precentral_L |
| **175** | 48 | 25 | 27 | Fronto-parietal Task Control | Frontal_Inf_Tri_R |
| **176** | -47 | 11 | 23 | Fronto-parietal Task Control | Frontal_Inf_Oper_L |
| **177** | -53 | -49 | 43 | Fronto-parietal Task Control | Parietal_Inf_L |
| **178** | -23 | 11 | 64 | Fronto-parietal Task Control | Frontal_Sup_L |
| **179** | 58 | -53 | -14 | Fronto-parietal Task Control | Temporal_Inf_R |
| **180** | 24 | 45 | -15 | Fronto-parietal Task Control | Frontal_Sup_Orb_R |
| **181** | 34 | 54 | -13 | Fronto-parietal Task Control | Frontal_Mid_Orb_R |
| **186** | 47 | 10 | 33 | Fronto-parietal Task Control | Frontal_Inf_Oper_R |
| **187** | -41 | 6 | 33 | Fronto-parietal Task Control | Precentral_L |
| **188** | -42 | 38 | 21 | Fronto-parietal Task Control | Frontal_Mid_L |
| **189** | 38 | 43 | 15 | Fronto-parietal Task Control | Frontal_Mid_R |
| **190** | 49 | -42 | 45 | Fronto-parietal Task Control | SupraMarginal_R |
| **191** | -28 | -58 | 48 | Fronto-parietal Task Control | Parietal_Sup_L |
| **192** | 44 | -53 | 47 | Fronto-parietal Task Control | Parietal_Inf_R |
| **193** | 32 | 14 | 56 | Fronto-parietal Task Control | Frontal_Mid_R |
| **194** | 37 | -65 | 40 | Fronto-parietal Task Control | Angular_R |
| **195** | -42 | -55 | 45 | Fronto-parietal Task Control | Parietal_Inf_L |
| **196** | 40 | 18 | 40 | Fronto-parietal Task Control | Frontal_Mid_R |
| **197** | -34 | 55 | 4 | Fronto-parietal Task Control | Frontal_Mid_L |
| **198** | -42 | 45 | -2 | Fronto-parietal Task Control | Frontal_Mid_Orb_L |
| **199** | 33 | -53 | 44 | Fronto-parietal Task Control | Parietal_Inf_R |
| **200** | 43 | 49 | -2 | Fronto-parietal Task Control | Frontal_Mid_Orb_R |
| **201** | -42 | 25 | 30 | Fronto-parietal Task Control | Frontal_Inf_Tri_L |
| **202** | -3 | 26 | 44 | Fronto-parietal Task Control | Frontal_Sup_Medial_L |
| **203** | 11 | -39 | 50 | Salience | Cingulum_Mid_R |
| **204** | 55 | -45 | 37 | Salience | SupraMarginal_R |
| **205** | 42 | 0 | 47 | Salience | Precentral_R |
| **206** | 31 | 33 | 26 | Salience | Frontal_Mid_R |
| **207** | 48 | 22 | 10 | Salience | Frontal_Inf_Tri_R |
| **208** | -35 | 20 | 0 | Salience | Insula_L |
| **209** | 36 | 22 | 3 | Salience | Insula_R |
| **210** | 37 | 32 | -2 | Salience | Frontal_Inf_Orb_R |
| **211** | 34 | 16 | -8 | Salience | Insula_R |
| **212** | -11 | 26 | 25 | Salience | Cingulum_Ant_L |
| **213** | -1 | 15 | 44 | Salience | Supp_Motor_Area_L |
| **214** | -28 | 52 | 21 | Salience | Frontal_Mid_L |
| **215** | 0 | 30 | 27 | Salience | Cingulum_Ant_L |
| **216** | 5 | 23 | 37 | Salience | Cingulum_Mid_R |
| **217** | 10 | 22 | 27 | Salience | Cingulum_Ant_R |
| **218** | 31 | 56 | 14 | Salience | Frontal_Sup_R |
| **219** | 26 | 50 | 27 | Salience | Frontal_Mid_R |
| **220** | -39 | 51 | 17 | Salience | Frontal_Mid_L |

---

**Supplemental Information Table 2.** ACT Intervention

**Weekly ACT Session Topics and Objectives:**
|  | **Session Topic** | **Session Objective** | **Homework Assignment** |
| --- | --- | --- | --- |
| **Week 1** | Introductions & Foundations of Treatment | Examine pain duration and analyze past treatment | Creative Hopelessness & Mindfulness<br>Practice: Monitor negative experiences/thoughts, and responses to those experiences throughout the week.<br>Complete 3 five-minute mindfulness practice sessions |
|  | Setting a Course for Treatment | Introduce behavioral model; in-vivo mindfulness practice | Check-in with yourself: Take 10-15 minutes each day to notice and observe. |
| **Week 2** | “Learning to Live” with Chronic Pain | Introduce/identify acceptance and values; in-vivo mindfulness practice | Values assessment: Identify personal values, rank importance and success |
|  | Values and Action | Values clarification and goal setting; identify barriers; in-vivo mindfulness practice | Goals and action: Identify values and goals consistent with each value |
| **Week 3** | Urges, Thoughts, & Feelings | Defusion of threats/barriers; in-vivo mindfulness practice | Observing your actions: Track actions and observations throughout the week |
|  | Action – Getting Your Feet Moving | Committed action; in-vivo mindfulness practice | Commit yourself to action: Track progress, difficulties, and experiences and review throughout the week |
| **Week 4** | Commitment | Acceptance, willingness, and barriers to willingness; in-vivo mindfulness practice | Mindfulness practice: Try new ways to practice mindfulness in daily life |
|  | Lifelong Maintenance | Values clarification; dealing with relapses and setbacks; review treatment plan | Lifelong assignment: Continued values and goals assessment; mindfulness practice |

## Description of the ACT therapist training process can be accessed at

Aytur, S., Campbell, C., Meier, S., et al. (2019). Broadening the scope of Acceptance and Commitment Therapy as a public health intervention for persons with chronic pain. Presented at the American Public Health Association annual conference, Philadelphia, PA, Nov. 5, 2019.

https://apha.confex.com/apha/2019/meetingapp.cgi/Paper/453678

**Supplemental Information Table 3.** Behavioral Assessments

| Battery/Assessment |
| --- |
| Center for Epidemiologic Studies Depression Scale (CES-D; Radloff, 1977) |
| Brief Pain Inventory (BPI; Cleeland & Ryan, 1994) |
| Acceptance & Action Questionnaire (AAQ-II; Bond et al., 2011) |
| Chronic Pain Acceptance Questionnaire (CPAQ; Vowles et al., 2011) |
| Patient-Reported Outcomes Measurement Information System (PROMIS ®; Cella et al., 2010): Pain Interference, Pain Intensity, Pain Behavior |
| Quality of Life in Neurological Disorders (Neuro QoL; Cella et al., 2012) - Short Forms: Cognitive Function, Satisfaction with Social Roles and Activities, Stigma, Sleep, Positive Affect and Well-Being, Fatigue, Emotional and Behavioral Dyscontrol, Depression, Anxiety, Ability to Participate in Social Roles and Activities, Communication |

## Notes

### Competing Interest Statement

The authors have declared no competing interest.

### Clinical Trial

NCT04502992

### Funding Statement

No external funding was received. The authors were funded by an internal University of New Hampshire Collaborative Research Excellence (CoRE) award.

### Author Declarations

The University of New Hampshire IRB provided approval for this study.

